# Short-term HPV detection dynamics across hormonal contraceptive methods in adolescent girls and young women: a secondary analysis of a randomized trial in South Africa

**DOI:** 10.1101/2025.01.14.25320519

**Authors:** Ramla F. Tanko, Ongeziwe Taku, Zizipho Z. A. Mbulawa, Keletso Phohlo, Iyaloo Konstantinus, Christina Balle, Tanya Pidwell, Anna-Ursula Happel, Katherine Gill, Linda-Gail Bekker, Heather B. Jaspan, Anna-Lise Williamson, Jo-Ann S. Passmore

**Author notes:** **Correspondence:** Professor Jo-Ann Passmore, Centre for Epidemic Response and Innovation (CERI), School of Data Science and Computational thinking, Tygerberg Campus, Francie van Zijl Drive, Tygerberg, 7505, South Africa.

## Abstract

**Background:** Human papillomavirus (HPV) is highly prevalent in adolescent girls and young women (AGYW) in sub-Saharan Africa. We evaluated short-term differences in HPV prevalence and early detection dynamics across hormonal contraceptive methods in AGYW.

**Methods:** Ninety-eight AGYW (15–19 years) were randomized to receive norethisterone enanthate (Net-EN), combined oral contraceptive pills (COCPs), or a combined contraceptive vaginal ring (CCVR). Cervical samples were collected at baseline and 16-weeks for HPV genotyping.

**Results:** Baseline HPV prevalence was high (93/98; 94.9%) and remained high at 16-weeks (87/97; 89.5%), with no significant differences between contraceptive methods. Longitudinal analyses (n=87 paired samples) showed no statistically significant differences in HPV acquisition, persistence, or clearance.

**Conclusions:** No significant differences were observed in HPV outcomes between hormonal contraceptive methods over 16-weeks. Findings should be interpreted as exploratory given the small sample size and short follow-up.

## INTRODUCTION

Human papillomavirus (HPV) is one of the most prevalent sexually transmitted infections (STIs) globally and is causally associated with cervical cancer (1,2). Adolescent girls and young women (AGYW) in sub-Saharan Africa bear a disproportionate burden of HPV infection (3,4). Hormonal contraceptives may influence HPV acquisition and clearance, potentially through effects on mucosal immunity, epithelial integrity, and the vaginal microbiome (5–11). However, evidence on this relationship remains inconsistent, and data comparing different contraceptive methods are limited. Findings on the association between hormonal contraceptive use and HPV outcomes have been heterogeneous. Some observational studies have reported reduced HPV clearance or increased persistence among users of combined oral contraceptive pills (COCPs; 5), whereas others have found no association or effects limited to prolonged duration of use (6,8). Differences in study design, population characteristics, duration of exposure, and adjustment for behavioural and biological confounders likely contribute to these inconsistencies. Importantly, a meta-analysis suggested that any association between hormonal contraceptive use and HPV outcomes may be duration-dependent, with longer-term use more consistently linked to altered HPV persistence and cervical cancer risk, while short-term effects remain less well defined (8).

Many existing studies have evaluated hormonal contraceptive exposure as a single category rather than comparing individual methods (6,8), despite evidence that different contraceptive formulations and delivery routes may have distinct effects on the cervicovaginal environment (10-12). Given that hormonal contraceptives differentially influence genital tract inflammation, epithelial barrier function, and the vaginal microbiome (10-12), method-specific effects on HPV acquisition and persistence are biologically plausible but remain insufficiently characterised. To our knowledge, no randomized studies have specifically examined short-term HPV detection dynamics across different hormonal contraceptive methods in AGYW, a population with a high burden of HPV infection and unique biological and behavioural risk profiles.

Risk of HPV acquisition and persistence is shaped by interacting biological and behavioural factors, including host immune status, co-infections with other sexually transmitted pathogens, and sexual practices such as early sexual debut and multiple partners (13-15). In this context, this study evaluated HPV prevalence and early detection dynamics over a 16-week randomized exposure period among South African AGYW using Net-EN, CCVR, or COCPs.

## METHODS AND MATERIALS

### Description of study cohort

This sub-study utilised data from the uChoose randomized crossover trial (NCT02404038) (16). Adolescent girls and young women (AGYW) aged 15–19 years were randomized to receive norethisterone enanthate (Net-EN), combined oral contraceptive pills (COCPs), or a combined contraceptive vaginal ring (CCVR) for 16 weeks. To avoid potential carryover effects, only data from the first randomized exposure period were included in this analysis. Cervical samples were collected at baseline and at 16 weeks for HPV genotyping. Although the parent study was a randomized controlled trial, this sub-study is observational in design and is reported in accordance with the STROBE guidelines for observational studies. As described previously, vulvovaginal swabs were screened for *Chlamydia trachomatis, Neisseria gonorrhoea, Trichomonas vaginalis*, and *Mycoplasma genitalium* by multiplex PCR, BV was diagnosed using Nugent scoring, and vaginal pH levels were measured using color-fixed indicator strips (Macherey-Nagel, Düren, Germany) (9-11, 17-18). Participants with STIs or BV Nugent scores 7-10 were managed according to national guidelines within the parent study. The parent uChoose study and this sub-study were approved by the University of Cape Town Human Research Ethics Committee (HREC Ref: 801/2014), and all participants provided written informed consent.

### HPV genotyping

HPV genotyping was performed at baseline and after 16 weeks on extracted DNA (MagNA Pure Compact Nucleic Acid Isolation Kit; Roche), from endocervical swabs collected during the speculum examination, using the HPV Direct Flow Chip test (Master Diagnóstica, Granada, Spain), according to the manufacturers’ instructions. The Direct Flow Chip detects 36 different HPV genotypes, including the 12 HR types based on recent IARC-aligned causal attribution analyses of HPV genotypes associated with invasive cervical cancer (2) (HPV-16, -18, -31, -33, -35, -39, -45, -51, -52, -56, -58, and -59). The assay also detects an additional 5 HPV types previously considered HR (26, 53, 66, 73, 82; according to the kit manufacturers), HPV-68, as well as 18 low-risk (LR) types (6, 11, 40, 42, 43, 44, 54, 55, 61, 62, 67, 69, 70, 71, 72, 81, 84, 89). For the purposes of HR-HPV analysis, HR-HPV types as defined by the IARC-aligned causal attribution framework (2); genotypes previously classified as HR but no longer included are shown separately. For the purposes of LR-HPV analysis, the manufacturers definition of LR-HPV types was used.

### Statistical analyses

HPV outcomes were defined as follows: (1) Acquisition - detection of a genotype at 16 weeks not present at baseline; (2) Persistence - detection of the same genotype at both time points; or (3) Clearance - absence at 16 weeks of a genotype detected at baseline. As HPV outcomes were evaluated only during the first randomized exposure period, analyses were conducted using a parallel-group approach based on initial contraceptive assignment. Cross-sectional analyses of HPV prevalence and genotype distribution at baseline and 16 weeks included all available samples at each time point, whereas longitudinal analyses of HPV acquisition, persistence, and clearance were restricted to participants with paired baseline and 16-week samples. Fisher’s exact test and descriptive statistics were used to compare differences between hormonal contraceptive arms for categorical variables. Changes between baseline and 16 weeks within the same contraceptive arm were assessed using McNemar’s test for paired binary data. Continuous variables were described using median and interquartile range (IQR) and compared using the Kruskal–Wallis test. Statistical analyses were performed using GraphPad Prism version 9 (GraphPad Software, USA). Given the exploratory nature of the study and limited statistical power, findings are interpreted cautiously and emphasis is placed on effect estimates and descriptive patterns rather than statistical significance.

### Trial registration

ClinicalTrials.gov NCT02404038. Registered 30 March 2015.

## RESULTS

HPV genotyping data were available for 98 participants at baseline and 95 at 16 weeks (Supplementary Figure 1), with 87 paired samples. Most participants reported prior hormonal contraceptive use at enrolment (Table 1), and all participants were HIV-negative. Despite this, the cohort reflected a high-risk behavioural and clinical context, including early sexual debut, multiple sexual partners, and a high burden of bacterial STIs and BV at baseline, which did not differ by study arm.

**Table 1.** Baseline demographic, behavioural, and clinical characteristics of adolescent girls and young women enrolled in the uChoose HPV sub-study.

| Contraceptive arm | All | Net-EN<sup>a</sup> | COCp | CCVR<sup>b</sup> | *P* |
| --- | --- | --- | --- | --- | --- |
| n | 98/130 (75.4%) | 44/98 (44.9%) | 24/98 (24.5%) | 25/98 (25.5%) |  |
| Age (years)* | 17.0 (16.0 – 18.0) | 17.0 (16.0 – 18.0) | 16.0 (16.0 – 18.0) | 17.0 (16.0 – 18.5) | 0.78 |
| Body mass index (kg/m<sup>2</sup>)* | 25.1 (21.5 – 29.1) | 25.8 (22.6 – 29.8) | 25.5 (20.7 – 30.5) | 24.9 (21.1 – 26.5) | 0.64 |
| Age of sexual debut (years)* | 13 (12.0 – 14.0) | 13 (12.0 – 14.0) | 14 (12.0 – 15.0) | 13 (12.0 – 15.0) | 0.32 |
| Condom use during last sex | 55/98 (56.1%) | 24/44 (54.5%) | 14/24 (58.3%) | 15/25 (60.0%) | 0.93 |
| >1 sex partners in past year | 21/98 (21.4%) | 10 (27.8%) | 5 (20.8%) | 6 (24.0%) | 0.79 |
| New partners in past year | 28/98 (28.6%) | 10 (22.7%) | 7 (29.2%) | 10 (40.0%) | 0.38 |
| Prior HC use: | 76/98 (77.6%) | 34/44 (77.3%) | 17/24 (70.8%) | 21/25 (84.0%) | 0.67 |
| DMPA | 9/98 (9.2%) | 4 (9.1%) | 4 (16.7%) | 1 (4.0%) |  |
| Implanon | 2/98 (2.0%) | 1(2.3%) | 0 (0.0%) | 1 (4.0%) |  |
| COCps | 3/98 (3.1%) | 2 (4.5%) | 0 (0.0%) | 1 (4.0%) |  |
| Net-EN | 62/98 (63.3%) | 27 (61.4%) | 13 (54.2%) | 18 (72.0%) |  |
| Vaginal pH* | 4.9 (4.4 – 5.3) | 5.0 (4.7 – 5.3) | 4.7 (4.4 – 5.0) | 4.7 (4.4 – 5.3) | 0.41 |
| Bacterial vaginosis: | 42/98 (42.9%) | 20/44 (45.5%) | 10/24 (41.7%) | 11/25 (44.0%) | 0.89 |
| STIs |  |  |  |  |  |
| *Chlamydia trachomatis* | 31/98 (31.6%) | 12/44 (27.3%) | 9/24 (37.5%) | 9/25 (36.0%) | 0.61 |
| *Neisseria gonorrhoeae* | 12/98 (12.2%) | 5/44 (11.4%) | 3/24 (12.5%) | 3/25 (12.0%) | 0.99 |
| *Trichomonas vaginalis* | 8/98 (8.2%) | 5/44 (11.4%) | 1/24 (4.2%) | 2/25 (8.0%) | 0.72 |
| *Mycoplasma genitalium* | 3/98 (3.1%) | 2/44 (4.6%) | 1/24 (4.2%) | 0/25 (0.0%) | 0.85 |

\*Median and 25<sup>th</sup> and 75<sup>th</sup> percentile; n: sample size
<sup>a</sup>No record available for age of sexual debut (n=3), condom use during last sex (n=8), numbers of sexual partners (n=8), new partners (n=9) and endogenous hormone levels (n=2) and n=1
<sup>b</sup>No data available for endogenous hormone levels (n=1)
Abbreviations: COCP, combined oral contraceptive pill; CCVR, combined contraceptive vaginal ring; Net-EN, norethisterone enanthate; DMPA, depot medroxyprogesterone acetate. No statistically significant differences were observed between arms at baseline.

Baseline HPV (93/98; 94.9%) and high-risk HPV (HR-HPV; 77/98; 78.6%) prevalence was high and did not differ by study arm (Table 2), with most AGYW having multiple HPV infections (79/98; 80.5%). HPV-31 and HPV-39 were the most frequently detected HR types (each 18/98; 18.4%), while HPV-16 and HPV-18 were detected in 6/98 (6.1%) and 14/98 (14.2%) participants, respectively (Supplementary Figure 2). HPV genotypes targeted by currently available bivalent and nonavalent vaccines accounted for a minority of detected high-risk HPV types, with non-vaccine genotypes predominating (Supplementary Figure 2).

**Table 2.**
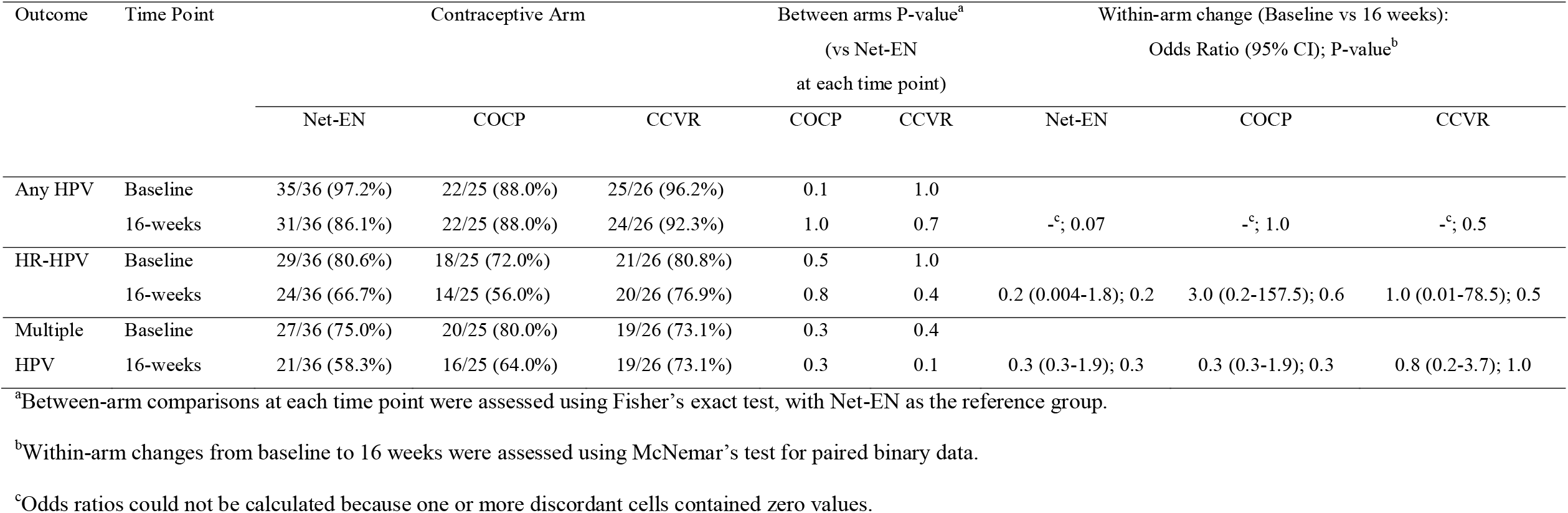
HPV prevalence in participants with paired baseline and week-16 samples.

At 16 weeks, HPV prevalence remained high (85/95; 89.5%), with no statistically significant differences between contraceptive methods (Table 2). Cross-sectional and longitudinal analyses yielded consistent findings, with no statistically significant differences in HPV outcomes observed across contraceptive methods. Longitudinal analyses further showed no significant differences in HPV acquisition, persistence, or clearance across arms (Supplementary Figure 3). Although no statistically significant differences were observed, descriptive patterns were noted across contraceptive methods. The combined prevalence of newly acquired and persistent HR-HPV genotypes was numerically higher among AGYW randomized to the CCVR arm, whereas a higher proportion of cleared HR-HPV genotypes was observed among those using Net-EN over the study period.

## DISCUSSION

This exploratory sub-study in the uCHOOSE randomized controlled trial found no significant differences in HPV prevalence or detection dynamics across hormonal contraceptive methods over 16 weeks, consistent with prior studies showing heterogeneous and often inconclusive associations (5-9). However, the descriptive patterns observed across contraceptive arms were biologically plausible and consistent with established effects of hormonal contraceptives on mucosal immunity, epithelial integrity, and the vaginal microbiome (10-12). This study should be interpreted as a short-term randomized exposure study designed to identify early biological signals in HPV detection patterns rather than to infer long-term infection dynamics.

The extremely high HPV prevalence (∼95%) observed in this cohort is consistent with previous studies in AGYW in sub-Saharan Africa (19) and likely reflects the high-risk behavioural and clinical context of the study population (13,16). Early sexual debut, multiple sexual partners, and inconsistent condom use are well-established risk factors for HPV acquisition (13) and were common in this cohort. Furthermore, the high prevalence of bacterial STIs and BV may have contributed to increased susceptibility to HPV acquisition and persistence through effects on genital tract inflammation and epithelial barrier disruption (10-12).

Although descriptive patterns in HPV clearance and acquisition were observed, these were not statistically significant. These observations align with known effects of hormonal contraceptives on the cervicovaginal environment (10-12). Previous work has demonstrated that Net-EN use is associated with increased microbial diversity and pro-inflammatory taxa, COCP use with *Lactobacillus*-dominant communities and reduced inflammation, and CCVR use with increased inflammatory signalling, including IL-1 and NF-κB pathways, alongside reduced epithelial barrier integrity (10-12). These distinct mucosal profiles may differentially influence HPV acquisition, persistence, and clearance. Genital inflammation may impair viral clearance and promote HPV persistence, while epithelial barrier disruption may increase susceptibility to HPV acquisition (20).

Evidence from studies of other contraceptive methods further supports the concept of method-specific effects on HPV outcomes. IUD use has generally not been associated with increased HPV acquisition, and some studies suggest enhanced clearance among copper IUD users, potentially through pro-inflammatory local immune responses (21-24). In contrast, hormonal IUDs may exert more local immunomodulatory effects on epithelial and cytokine profiles, with variable associations reported (24,25). These findings reinforce that contraceptive-associated effects on HPV dynamics are likely mediated through distinct influences on mucosal immunity and epithelial integrity.

This study has several limitations. First, this study was not powered to detect modest differences between contraceptive methods. Second, the 16-week follow-up period is insufficient to assess true HPV persistence or clearance, which typically occur over months to years (26). Third, the absence of a non-hormonal contraceptive control group limits causal inference, and the findings reflect comparisons between methods rather than effects of hormonal contraception. Fourth, most participants had prior hormonal contraceptive exposure, limiting assessment of first-time exposure. The extremely high baseline HPV prevalence (∼95%) limits the ability to detect incident infections and may bias estimates of acquisition and clearance. Finally, residual confounding by factors such as HPV vaccination, smoking, and sexual behaviour cannot be excluded (13).

These findings highlight the importance of considering contraceptive-specific effects on the genital mucosal environment when evaluating susceptibility to HPV and other STIs. Although no statistically significant differences were observed, descriptive patterns were consistent with biologically plausible mechanisms. Further studies are needed to determine whether these differences translate into clinically meaningful effects, with implications for contraceptive counselling and cervical cancer prevention strategies in high-burden settings.

## Data Availability

The datasets generated during the current study are not publicly available due to ethical and participant confidentiality restrictions but are available from the corresponding author on reasonable request and subject to approval by the University of Cape Town Human Research Ethics Committee.

## SUPPLEMENTARY INFORMATION

Supplementary information accompanies this paper.

## DECLARATIONS

### ETHICS APPROVAL AND CONSENT TO PARTICIPATE

The parent uChoose study and this HPV sub-study were conducted in accordance with the Declaration of Helsinki and Good Clinical Practice guidelines. Ethical approval was obtained from the University of Cape Town Human Research Ethics Committee (HREC Ref: 801/2014). Written informed consent was obtained from all participants prior to enrolment. For participants younger than 18 years, written informed assent was obtained alongside parental or legal guardian consent, in accordance with local regulatory requirements. Participants were informed of the study procedures, potential risks and benefits, and their right to withdraw at any time without penalty. All samples and data used in this sub-study were collected as part of the approved parent study protocol. The current analysis utilised de-identified data, and no additional participant contact or procedures were required. All participant data were anonymised prior to analysis, and confidentiality was maintained throughout the study.

### CONSENT FOR PUBLICATION

Not applicable.

### AUTHOR CONTRIBUTIONS

Conceived and designed the experiments: J.A.P, R.F.T., H.J., A.L.W. Designed and recruited the study participants: L.G.B., K.G., T.P., and S.B. Processed samples: I.K., C.B., S.J., A.H., and H.G. Performed the experiments: T.F.M., K.P., O.T., and Z.Z.A.M. Analysed the data: R.F.T., and J.A.P. Wrote the manuscript: All authors. Patients and/or the public were not involved in the design, conduct, reporting, or dissemination plans of this research.

## ACKNOWLEDGMENTS

We are grateful to all the uChoose study participants and protocol team: Ms Penelope Ngcobo for recruiting and consenting participants, Dr Eve Mendel the study clinician, Sister Janine Nixon the study nurse and Ms Keshani Naidoo for processing the samples, Ms Maria Mwakatima and Zoe Baker for editorial and review support.

## SUPPLEMENTARY FIGURE LEGENDS

**Supplemental Figure 1.**
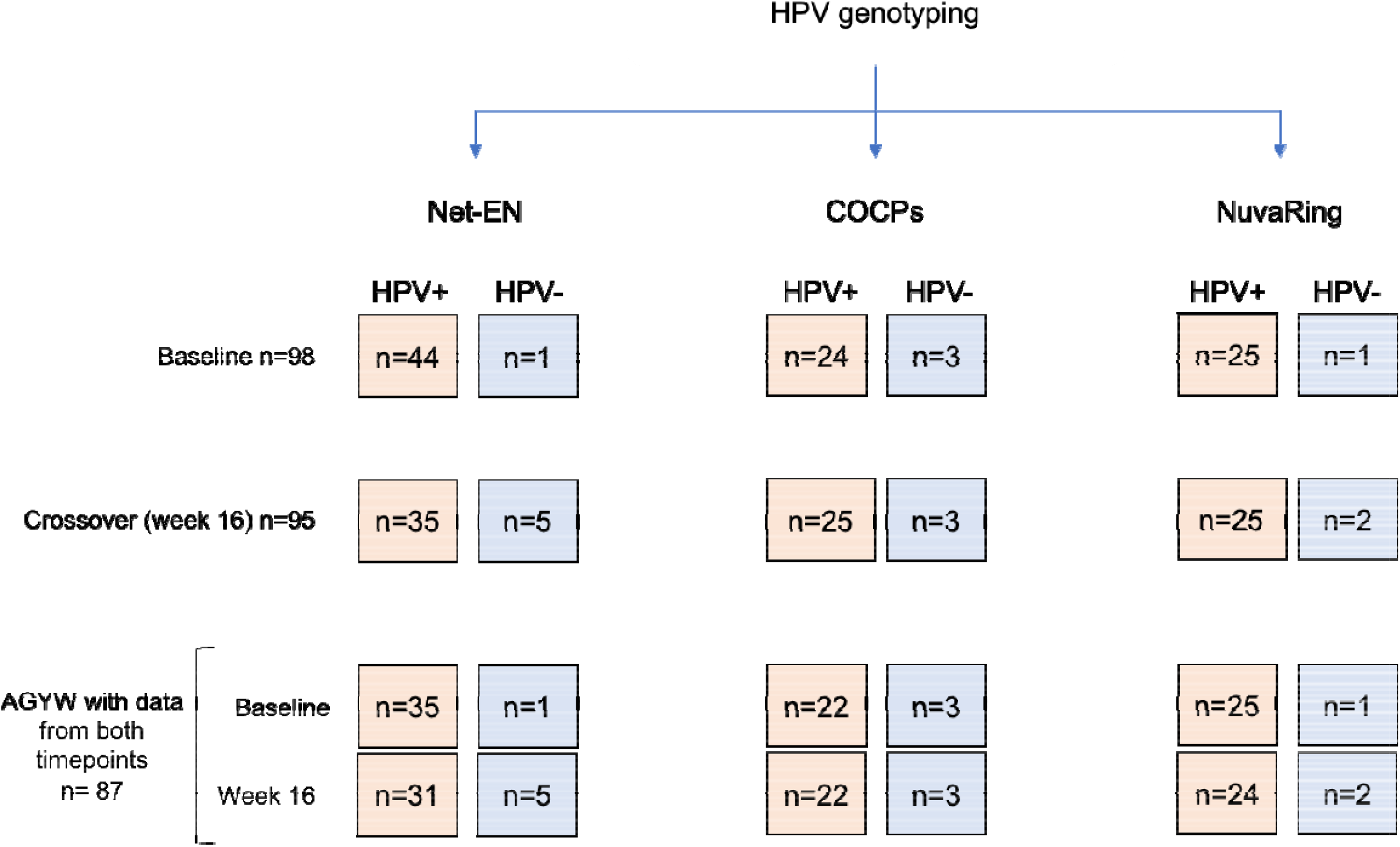
Study design overview. Study design for the open-label, randomized crossover uChoose trial conducted among adolescent girls and young women (AGYW). Participants were randomized to one of three hormonal contraceptive methods: combined oral contraceptive pills (COCPs), injectable norethisterone enanthate (Net-EN), or the combined contraceptive vaginal ring (CCVR). In the parent uChoose trial, participants used an assigned hormonal contraceptive method for 16 weeks before crossing over to an alternative method, with no washout period between interventions; the primary outcomes of the parent trial were acceptability, feasibility, and adherence. The present HPV sub-study was restricted to samples collected at baseline and after the initial 16-week randomized exposure period, prior to crossover, and evaluated HPV prevalence and genotype dynamics. Accordingly, analyses presented here reflect comparisons across contraceptive methods during the first exposure period only. Numbers (n) indicate the number of participants with available and typeable HPV genotyping data for the specific outcome and time point shown; variation in n across contraceptive arms reflects missing or insufficient samples for genotyping at specific visits.

**Supplementary Figure 2.**
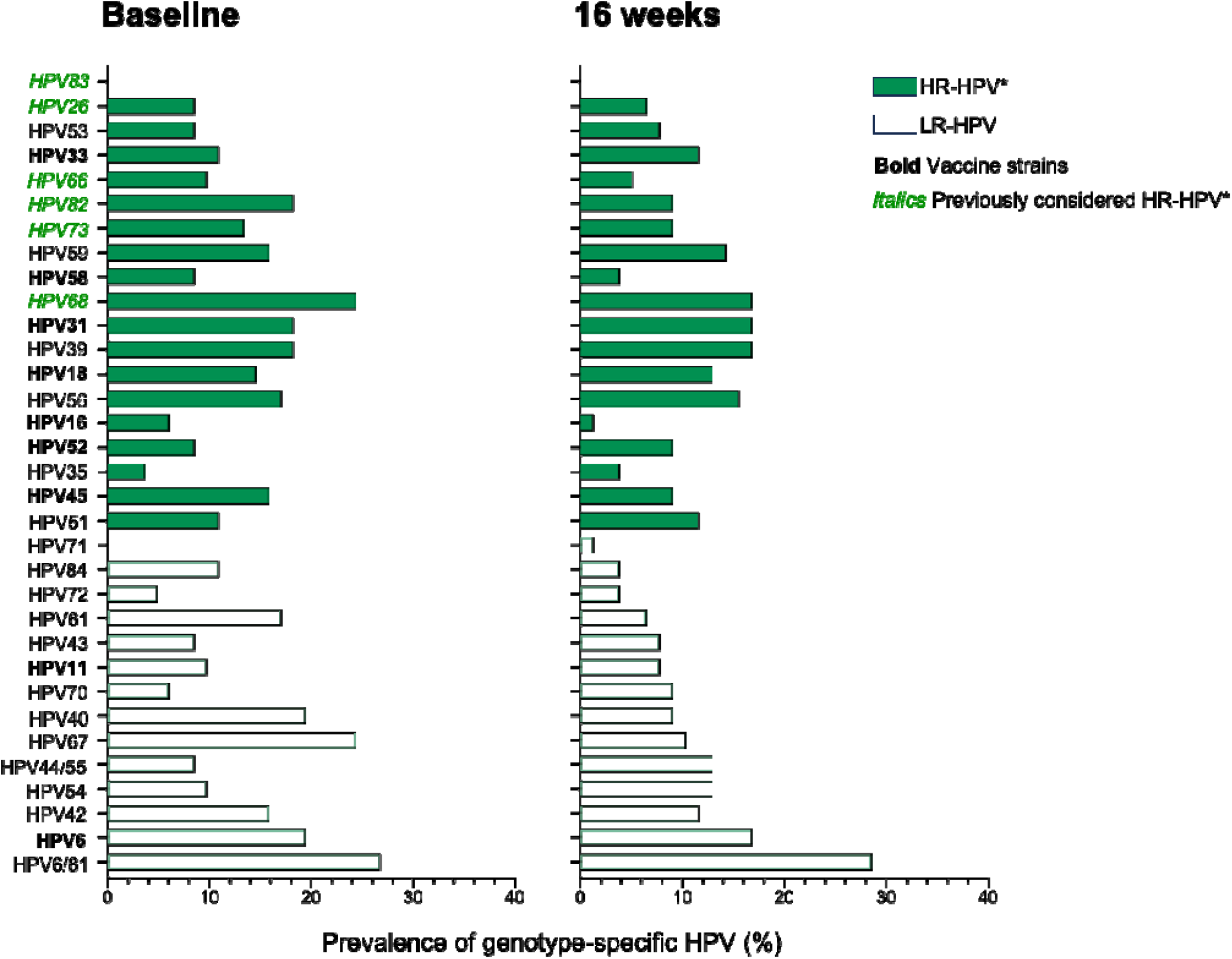
Distribution of HPV genotypes at baseline and 16 weeks by randomized contraceptive arm. Distribution of HPV genotypes among adolescent girls and young women (AGYW) at baseline and at 16 weeks, using all available samples at each time point. Each bar graph represents the prevalence of a specific HPV genotype. Light colours indicate low-risk HPV (LR-HPV) types, while dark colours represent high-risk HPV (HR-HPV) types. Genotypes no longer classified as high-risk by the International Agency for Research on Cancer (IARC) are shown in green italics. Genotypes targeted by the bivalent (HPV-16, -18), quadrivalent (HPV-6, -11, - 16, -18), and nonavalent (HPV-6, -11, -16, -18, -31, -33, -45, -52, -58) vaccines are highlighted in bold. Although the parent uChoose trial employed a crossover design, the present HPV sub-study includes only data from the first randomized exposure period (baseline to 16 weeks), prior to crossover.

**Supplementary Figure 3.**
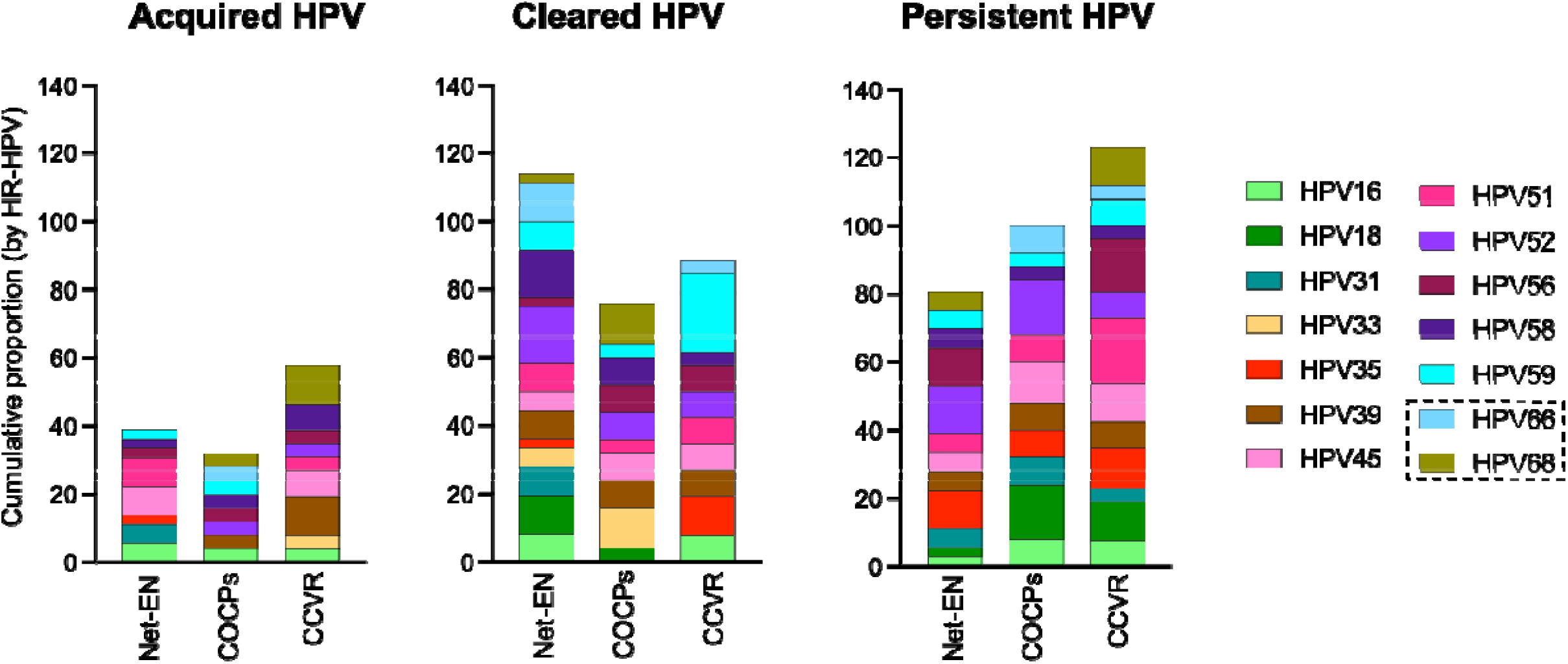
HPV Genotype Distribution Across Contraceptive Arms. Stacked bars show the prevalence of HR HPV types (including HPV-66 and -68 which were previously classified as HR) that were newly acquired between 0 and 16 weeks (left panel), cleared (middle panel), or persisted (right panel), stratified by genotype and contraceptive method, based on paired baseline and week 16 samples only. Panels represent data for Net-EN (n=36), COCPs (n=25), and CCVR (n=26), based on paired samples between baseline and week 16 visits.

